# Outcomes of the three-month weekly isoniazid with rifapentine (3HP) versus the six-month isoniazid preventive therapy (6H) among people newly enrolled in HIV care in western Kenya

**DOI:** 10.64898/2026.03.04.26347621

**Authors:** Dickens O. Onyango, Jerphason O. Mecha, Lilian N. Njagi, Samson Aoro, Timothy Malika, John Kinuthia, Grace John-Stewart, Sylvia M. LaCourse

**Affiliations:** Kisumu County Department of Health, Kisumu, Kenya; Department of Research and Programs, Kenyatta National Hospital, Nairobi, Kenya; Centre for Respiratory Disease Research, Kenya Medical Research Institute, Nairobi, Kenya; Department of Global Health, University of Washington, Seattle, Washington, USA; Department of Medicine, Division of Allergy and Infectious Diseases, University of Washington, Seattle, Washington, USA; Department of Epidemiology, University of Washington, Seattle, Washington, USA; Department of Pediatrics, University of Washington, Seattle, Washington, USA

**Author notes:** **Corresponding author:** Dr. Dickens Onyango, Kisumu County Department of Health, Kisumu Kenya, P.O. BOX 3670-40100, Kisumu. **Funding:** This research was funded by the Firland Foundation under award number 20220018, a 2023 developmental grant from the University of Washington / Fred Hutch Center for AIDS Research, an NIH funded program under award number AI027757, and the Seattle Tuberculosis Research Advancement Center (SEATRAC), an NIH-funded program under award number P30AI168034-01. The content is solely the responsibility of the authors and does not necessarily represent the official views of the Firland Foundation and the NIH. **Conference presentation:** Preliminary data were presented at the 32nd Conference on Retroviruses and Opportunistic Infections (CROI 2025), **9-12 March 2025**, San Francisco, USA.

**Keywords:** Human immunodeficiency virus, Mycobacterium tuberculosis infection, preventive therapy, isoniazid, rifapentine

## Abstract

**Background:** In trials, three-month weekly rifapentine and isoniazid (3HP) showed higher adherence and completion than the six-month daily isoniazid (6H) regimen for TB preventive treatment (TPT). However, programmatic outcome data remain limited.

**Methodology:** We evaluated the TPT cascade among people with HIV (PWH) aged ≥15 years newly enrolled in HIV care in western Kenya. Initiation and completion of 6H (Jan to Sept 2022) were compared to 3HP (Oct 2022-Sept 2023) using Chi-square tests. Correlates of non-initiation and non-completion were assessed using Poisson regression with generalized linear models. Mortality within 24 months was evaluated using Cox proportional hazards regression.

**Results:** Of 1,930 PWH (median age 33 years [IQR=27-41]), 65.8% were female, and 19.5% had AHD at enrolment. Overall, 1,922 (99.6%) were screened for active TB, of whom 1,790 (97.5%) were TPT-eligible; 1577 (88.1%) of these initiated TPT. TPT initiation was higher with 3HP than 6H (89.8% vs. 84.2%; p<0.001). TPT completion was similar for 3HP and 6H (89.2% vs. 88.8% p=0.77). TB incidence (per 1,000 person-months) was lower among TPT-completers (0.22; 95% CI 0.15-0.35) than those who neither initiated (4.25; 95% CI 1.77-10.23) nor completed TPT (3.75; 95% CI 2.49-5.64). AHD was associated with higher risk of TPT non-initiation (aRR=2.14; 95% CI 1.59-2.87) and non-completion of both 6H (aRR=2.56; 95% CI: 1.55-4.23) and 3HP (aRR=1.68; 95% CI 1.07-2.63).

**Conclusions:** 3HP increased TPT initiation but did not improve completion rates compared to 6H. Targeted interventions are needed to support 3HP completion, particularly in PWH with AHD

**Key points:** We compared 3HP and 6H for TB prevention in people with HIV in western Kenya. 3HP led to better initiation and both had high completion rates. Advanced HIV disease affected participation and non-completers faced significant mortality.

## INTRODUCTION

A quarter of the world’s population are infected with *Mycobacterium tuberculosis (MTb)*, with a 5-10% risk of developing TB disease in their lifetime (1). The risk of TB disease in people with HIV (PWH) with MTb infection is 7-10% annually and 30% over their lifetime (2, 3). TPT reduces the risk of TB by 60% and lowers mortality by 39%, especially when combined with antiretroviral therapy (ART) (3-5). TPT with six months of daily isoniazid (6H) or alternative regimens is recommended for PWH aged at least 12 months in settings with high TB prevalence (6), but its uptake and completion remain low, especially in low- and middle-income countries (LMICs) (7, 8). The underutilization of TPT in LMICs has been attributed to commodity stock-outs, lack of health provider buy-in, and long duration of 6H (9). However, factors associated with completion of shorter isoniazid-rifapentine regimens have not been documented.

Programmatic implementation of TPT involves a complex cascade with multiple steps, including eligibility screening, initiation, follow-up, and completion, with drop-offs at each step undermining TPT’s effectiveness (10). According to the World Health Organization (WHO), in 2024, the global TPT coverage among PWH was estimated at 58% against a target of over 90% (11). To improve TPT uptake and completion, WHO recommended the 3-month course of weekly isoniazid (H) with rifapentine (P) (3HP) for PWH in 2018 (12). Although clinical trials showed higher uptake, adherence, and completion rates with 3HP (13, 14), real-world data remain limited, especially among PWH with advanced HIV disease (AHD). Kenya adopted WHO guidelines and rolled out 3HP among PWH aged ≥15 in 2022 (15). Since then, the 3HP cascade has not been evaluated, and factors associated with its non-completion remain unknown.

We conducted a retrospective review of the medical charts of PWH who enrolled in care before and after the implementation of the 3HP regimen in western Kenya. We compared the completion of the TPT cascade for 6H versus 3HP. We evaluated the correlates of TPT non-initiation, non-completion of 3HP and 6H, and mortality within 24 months of enrolment in HIV care.

## METHODS

### Study Setting

This study was conducted in seven high-volume HIV clinics in Kisumu County in western Kenya, with an HIV prevalence of 17.5% (16) and an annual TB case notification rate of 243 per 100,000 population (17). TPT services in the region follow national TPT guidelines. Prior to 2021, 6H was the primary regimen for all TPT-eligible populations, including PWH. In 2021, the guidelines were updated to recommend 3HP as the preferred TPT regimen in PWH aged ≥15 years, with programmatic rollout in 2022 (18). Newly enrolled PWH undergo WHO staging and are initiated on ART. CD4 count is recommended as a baseline investigation for all PWH, patients suspected of HIV treatment failure, and patients returning after interruption of care >3 months (19). PWH in WHO stage 3 or 4, or with a CD4 count <200 cells/µ, are considered as having AHD. Viral load testing is offered at month 3 for all PWH, then every 6 months for patients aged ≤24 years and annually for those aged ≥25 years. PWH are screened for TB disease using an intensive case finding (ICF) algorithm, which assesses cough (any duration), fever, night sweats, and weight loss. PWH without TB-suggestive symptoms are eligible for TPT, while those with TB-suggestive symptoms undergo diagnostic testing and receive TPT only if TB disease is ruled out (18). PWH who initiate TPT return for refills at intervals determined by their HIV clinics, typically aligned with ART visits. TPT data are recorded in hard copy HIV treatment registers or electronic medical records by nurses or clinicians.

### Study Design

We evaluated the TPT cascade using retrospective data collected with a standard electronic data abstraction tool programmed in REDCap and administered on tablet devices. Data of all PWH aged ≥15 years newly enrolled in HIV care from January 2022 through September 2023 were abstracted from the medical records and TPT registers. Socio-demographic information (age, sex, level of education, and occupation), screening for TPT eligibility, TPT regimen, clinical status at TPT initiation (WHO stage, CD4 count, viral load, presence of opportunistic infections), date of TPT initiation, and outcome of TPT (completion, loss to follow up, discontinuation/reason for discontinuation, TB diagnosis, death or transfer out) were abstracted. PWH who transferred out, discontinued their regimens, were lost to follow-up, or died before completion were classified as non-completers.

### Data analysis

We evaluated the TPT cascade among PWH entering HIV care from January to September 2022, when only 6H was implemented, and October 2022 to September 2023, when 3HP was the preferred regimen in Kenya. We compared the proportions of PWH who completed steps of the cascade (eligibility screening, initiation, and completion) for 6H and 3HP using Chi-square tests. PWH were classified as virally suppressed if their viral load was <200 HIV RNA copies per milliliter. Poisson regression with general linear models was used to assess the correlates of non-initiation of TPT and non-completion, with non-completion stratified by regimen (6H versus 3HP). We conducted survival analysis using all-cause mortality within 24 months of HIV care enrollment as the primary outcome, with time-to-event defined as the interval between enrolment and death. PWH who were lost to follow-up or transferred out were censored. Crude mortality rate (CMR) was calculated as the number of deaths per 100 person-years of observation, stratified by age group, sex, baseline WHO stage, and TPT status (ineligible [TB disease at enrolment], eligible but not initiated TPT, initiated TPT but not completed, and completed TPT). We evaluated the predictors of death within 24 months of enrolment using Cox proportional hazards regression models. Variables with a p-value of ≤0.1 during bivariate analysis were included in multivariate analysis. Factors with a p-value of <0.05 during multivariate analysis were considered statistically significant.

### Ethical considerations

Ethical approval for the study was granted by the Kenyatta National Hospital/University of Nairobi Ethics Review Committee and the Jaramogi Oginga Odinga Teaching and Referral Hospital Ethics Review Committee. Informed consent was waived as the study involved only the abstraction and analysis of data collected during routine clinical care.

## RESULTS

### Demographic and clinical characteristics of participants

A total of 1,930 PWH aged 15 years and above were newly enrolled in HIV care between January 2022 and June 2023. Their median age was 33 (interquartile range [IQR]=27-41) years (**Table 1**). Baseline WHO stage was available for 99.4% (n = 1, 920) of PWH, of whom 19.5% (n=374) were in WHO stage 3 or 4. CD4 counts were available for 56.8% (n=1096) of whom 30.9% (n=339) had <200 cells per cubic milliliter. Viral loads conducted within 6 months of enrollment in care were available for 56.7% (n = 1,095) of PWH, of whom 91.5% (n = 1,002) were suppressed.

**Table 1:** Demographic and clinical characteristics of people with HIV who newly initiated HIV care Jan 2022 to September 2023 in western Kenya (N=1, 930)

|  | All | 6H only period | 3HP period |
| --- | --- | --- | --- |
| Characteristic | Number (%) |  |  |
| **Age at enrollment** |  |  |  |
| Median age in years (interquartile range) | 33 (27-41) | 34 (27-41) | 34 (27-40) |
| 15 - 24 years | 320 (16.6) | 104 (14.5) | 216 (17.8) |
| 25 - 34 years | 723 (37.5) | 273 (38.1) | 450 (37.1) |
| 35 - 44 years | 540 (28.0) | 201 (28.1) | 339 (27.9) |
| 45 – 54 years | 243 (12.6) | 88 (12.3) | 155 (12.8) |
| 55+ | 104 (5.4) | 50 (7.0) | 54 (4.5) |
| **Sex** |  |  |  |
| Male | 661 (34.2) | 228 (31.8) | 433 (35.7) |
| Female | 1269 (65.8) | 488 (68.2) | 781 (64.3) |
| **Employment** |  |  |  |
| Formal employment | 236 (13.6) | 60 (9.9) | 176 (15.5) |
| Informal employment | 1120 (64.3) | 405 (67.1) | 715 (62.9) |
| Unemployed | 297 (17.1) | 115 (19.0) | 182 (16.0) |
| Student | 88 (5.1) | 24 (4.0) | 64 (5.6) |
| **Highest level of education** |  |  |  |
| Primary and below | 503 (30.0) | 201 (36.0) | 302 (27.1) |
| Secondary and above | 1171 (70.0) | 358 (64.0) | 813 (72.9) |
| **Marital status** |  |  |  |
| Married | 1140 (61.4) | 400 (59.8) | 740 (62.3) |
| Single | 436 (23.5) | 157 (23.5) | 279 (23.5) |
| Widowed/divorced | 280 (15.1) | 112 (16.7) | 168 (14.2) |
| **Facility type** |  |  |  |
| Referral Hospitals | 890 (46.1) | 339 (47.3) | 551 (45.4) |
| Non-referral hospitals | 1040 (53.9) | 377 (52.7) | 633 (54.6) |
| **WHO stage at enrolment** |  |  |  |
| Stage 1 | 1267 (66.0) | 495 (69.7) | 772 (63.8) |
| Stage 2 | 279 (14.5) | 94 (13.2) | 182 (15.3) |
| Stage 3/4 | 374 (19.5) | 121 (17.0) | 253 (20.9) |
| **Viral load (N=1095)** |  |  |  |
| Median (IQR) | 50 (0-50) | 50 (0-50) | 50 (0-50) |
| Suppressed | 1002 (91.5) | 294 (90.5) | 706 (92.0) |
| Non-suppressed | 93 (8.5) | 31 (9.5) | 62 (8.1) |

### TPT Cascade Performance: 3HP versus 6H

Over 99% (n = 1922) of PWH newly enrolled in HIV care were screened for TPT eligibility using the ICF tool (**Figure 1**). Overall, 16.5% (n = 317) of those who were screened had TB-suggestive symptoms and underwent evaluation for TB disease, including 8.9% (n = 172) with cough, 4.4% (n = 86), 3.7% (n = 71) with fever, and 3.3% (n = 64) with weight loss. The frequency of TB symptoms was higher in PWH with AHD (37.9%; 95% CI 33.0%-43.0%) than those without AHD (2.8%; 95% CI 2.1%-3.8%) [p-value <0.001]. TB disease was diagnosed in 6.9% (n = 132) of screened PWH (41.6% of those with TB-suggestive symptoms); 88.6% were microbiologically confirmed. Among PWH with TB-suggestive symptoms, prevalent TB disease was more common in those with AHD than those without AHD (85.8%; 95% CI 78.9%-91.1% vs. 22.7%; 95% CI 11.5%-37.8%; p-value <0.001).

**Figure 1:**
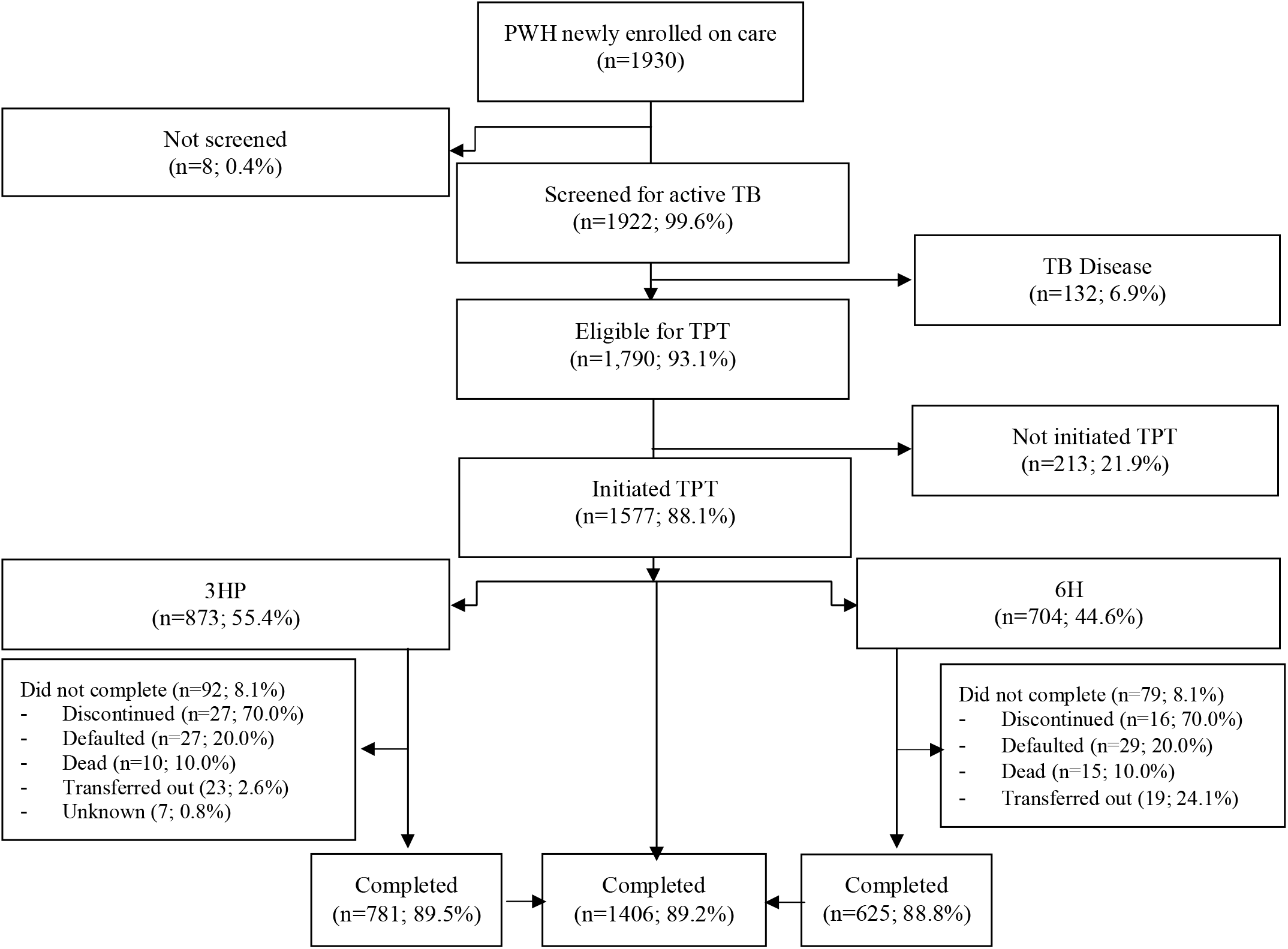
Overview of the TPT cascade of PWH who were enrolled in from January 2022 through September 2023 in western Kenya.

A total of 1,790 PWH (93.1% of those screened) were eligible for TPT, 88.1% (n = 1,577) of whom initiated TPT, with 55.4% (n = 873) initiating 3HP and 44.6% (n= 704) initiating 6H. TPT initiation was significantly higher during the 3HP period than the 6H period (89.8% vs. 84.2%; p<0.001) (**Figure S1**). However, 3HP completion was similar to 6H (89.2% vs. 88.8%, p=0.77). Discontinuation of 3HP due to adverse effects occurred rarely and was similar to 6H (0.7% [n = 6] vs. 0.6% [n = 4], p=0.76).

Among the 1,790 PWH who were TB free at enrolment, 2.6% (n=47) developed TB disease within 24 months of enrolment. TB incidence was substantially lower among participants who completed TPT (0.22 per 1,000 person-months; 95% CI 0.15-0.35) compared with those who never initiated (4.25 per 1,000 person-months; 95% CI 1.77-10.23) or did not complete treatment (3.75 per 1,000 person-months; 95% CI 2.49-5.64).

### Factors associated with TPT non-initiation and non-completion of 3HP and 6H

In multivariate analysis, TPT non-initiation was nearly three times higher among patients with primary level of education (aRR=2.98; 95% CI 1.85-4.81) than those with tertiary education (**Table 2**). Similarly, TPT non-initiation was 52% higher among students or unemployed PWH (aRR=1.52; 95% CI 1.18-1.96) than those with formal or informal employment. Receiving HIV care in referral hospitals was associated with a 94% (aRR=1.94; 95% CI 1.47-2.57) higher likelihood of not initiating TPT compared to receiving care in non-referral hospitals. Additionally, having AHD at enrolment was associated with a two-fold likelihood (aRR=2.14; 95% CI 1.59-2.87) of TPT non-initiation than those enrolled in WHO stage 1 or 2. Similarly, AHD at enrolment was associated with a higher risk of non-completion of both 6H (aRR=2.56; 1.55-4.23) and 3HP (aRR=1.68; 95% CI 1.07-2.63) (**Table 3**). Among PWH with AHD, TPT non-completion was non-significantly higher with 6H than 3HP (27%, 95% CI 18%-39% vs. 18%, 95% CI 12%-25%; aRR = 1.51; 95% CI: 0.89–2.53; p = 0.12). Being widowed or divorced was also associated with a higher risk of non-completion of 6H (aRR=1.98; 95% CI 1.19-3.27) and 3HP (aRR=1.58; 95% CI 1.01-2.50) compared to being married. Additionally, receiving HIV care in referral hospitals was associated with 6H non-completion (aRR=1.69; 95% CI 1.02-2.81) (**Table S1**).

**Table 2:**
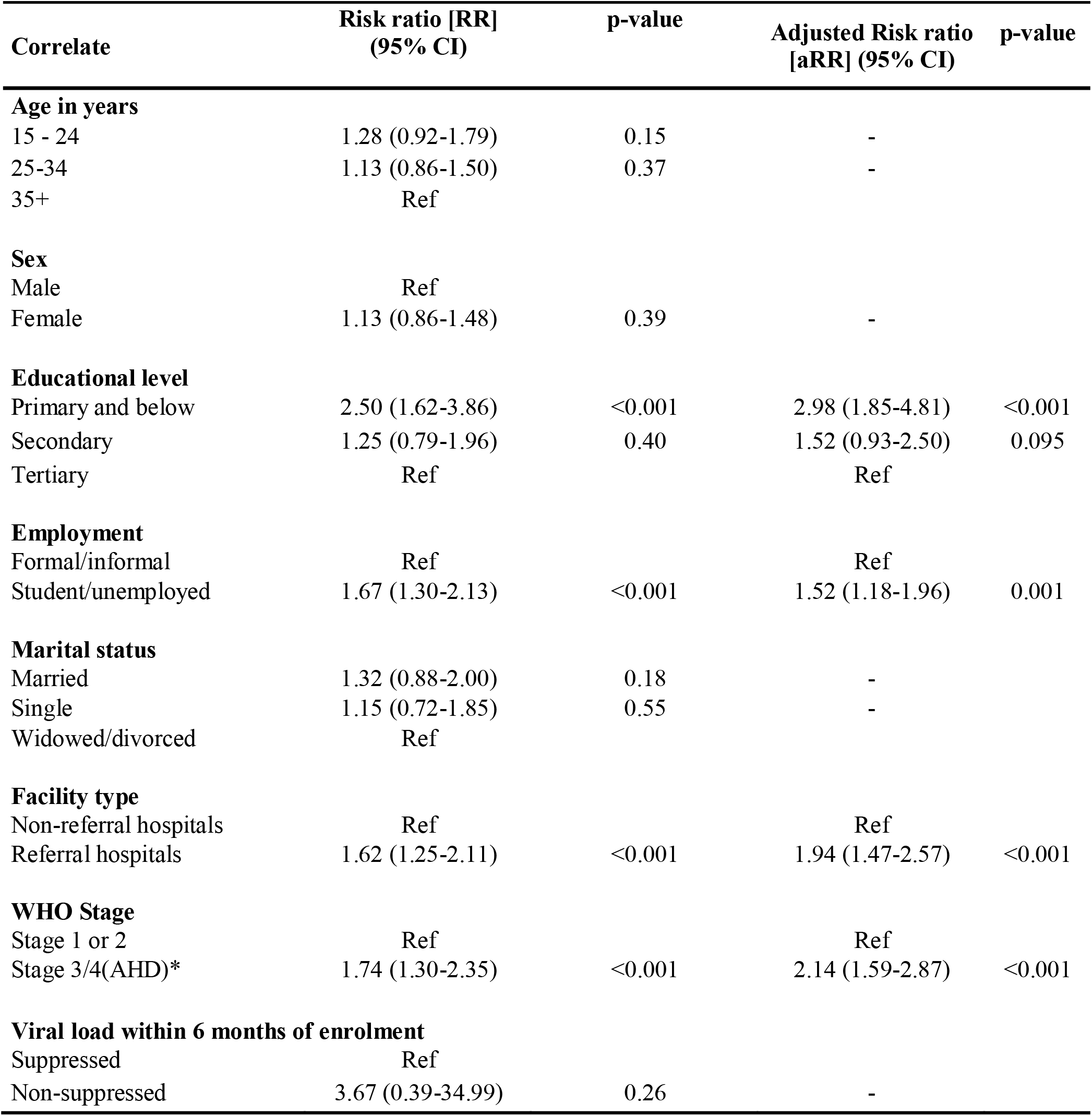
Correlates of non-initiation of TPT among PWH who were newly enrolled in HIV care from January 2022 through September 2023, western Kenya (N=1, 790)

**Table 3:** Correlates of 6H and 3HP non-completion among PWH who were newly enrolled in HIV care from January 2022 through September 2023, western Kenya (N=1,577)

| **Correlate** | **6H (N=704)** |  |  | **3HP (N=873)** |  |  |
| --- | --- | --- | --- | --- | --- | --- |
|  | Risk ratio [RR]<br>(95% CI) | Adjusted risk<br>ratio[aRR]*<br>(95% CI) | p-value | Risk ratio [RR]<br>(95% CI) | Adjusted risk<br>ratio [aRR]*<br>(95% CI) | p-value |
| **Age in years** |  |  |  |  |  |  |
| 15 - 24 | Ref |  |  | Ref |  |  |
| 25-34 | 0.83(0.46-1.49) | - |  | 0.70(0.41-1.20) | - |  |
| 35+ | 1.01(0.58-1.76) | - |  | 0.76(0.45- 1.26) | - |  |
| **Sex** |  |  |  |  |  |  |
| Male | Ref |  |  |  |  |  |
| Female | 1.02(0.64-1.62) | - |  | 0.84(0.57-1.25) | - |  |
| **Educational level** |  |  |  |  |  |  |
| Primary and below | 1.95(0.90-4.21) | 1.69(0.77-3.68) | 0.19 | 0.79(0.46-1.36) | 1.18(0.71- 1.94) | 0.53 |
| Secondary | 1.63(0.78-3.43) | 1.54(0.75- 3.15) | 0.24 | 0.85(0.54-1.35) | 1.45(0.83- 2.52) | 0.19 |
| Tertiary | Ref | Ref |  | Ref | Ref |  |
| **Employment** |  |  |  |  |  |  |
| Formal/informal | Ref |  |  | Ref |  |  |
| Student/unemployed | 0.76(0.47-1.25) | - |  | 0.90(0.58- 1.40) | - |  |
| **Marital status** |  |  |  |  |  |  |
| Married | Ref | Ref |  | Ref | Ref |  |
| Single | 0.95(0.54-1.67) | 0.91(0.52- 1.59) | 0.75 | 1.19(0.75- 1.90) | 1.29(0.81- 2.07) | 0.28 |
| Widowed/divorced | 1.93(1.17-3.17) | 1.98(1.19- 3.27) | 0.04 | 1.68(1.04-2.69) | 1.58(1.01- 2.50) | 0.04 |
| **Facility type** |  |  |  |  |  |  |
| Nonreferral hospitals | Ref | Ref |  | Ref |  |  |
| Referral hospitals | 1.93(1.24-2.98) | 1.69(1.02- 2.81) | 0.02 | 1.52(1.03- 2.23) | 1.36(0.88- 2.11) | 0.16 |
| **WHO Stage** |  |  |  |  |  |  |
| Stage 1 or 2 | Ref | Ref |  | Ref | Ref |  |
| Stage 3/4(AHD)* | 2.93(1.88-4.58) | 2.56(1.55- 4.23) | <0.001 | 1.92(1.26- 2.94) | 1.68(1.07- 2.63) | 0.02 |
| **Viral load** |  |  |  |  |  |  |
| Suppressed | Ref |  |  | Ref |  |  |
| Non-suppressed | 0.51(.070-3.71) | - |  | 0.90(0.22- 3.71) | - |  |

### Mortality within 24 months of enrolment in HIV care

Of the 1930 PWH enrolled in HIV care in the studied period, 80.1% (n=1,545) remained active on ART 24 months post enrollment, while 8.3% (n = 161) were lost to follow-up, 6.1% (n = 118) had transferred care out of the study area, and 5.6% (n = 108) had died, with a median time to death of 1.72 months (IQR: 0.51-3.66) (**Figure 2**). The crude mortality rate (CMR) was 5 (95% CI 4.1 – 6.0) per 100 person-years of observation (**Table S2**). The CMR among PWH who initiated but did not complete TPT (CMR=16.0; 95% CI 10.9 – 23.3) was as similar to those who were eligible but did not initiate TPT (CMR=18.8; 95% CI 14.0 – 25.3). In bivariate analysis, the risk of death was significantly higher among PWH who were aged 35 years and over at enrolment, males, those receiving care in referral hospitals, and with baseline WHO stages 2, 3, and 4 (**Table 4**). Study period and TPT regimen were not significantly associated with mortality. In multivariate analysis, PWH aged ≥35 years had a 94% higher likelihood of mortality within 24 months of enrolment than those aged 25–34 years (adjusted hazard ratio [aHR] = 1.94; 95% CI: 1.38–2.75). Mortality risk rose progressively with advancing WHO stage at enrolment: stage 2 (aHR = 5.95; 95% CI: 2.03– 17.48), stage 3 (aHR = 7.70; 95% CI: 3.25–18.20), and stage 4 (aHR = 21.77; 95% CI: 8.17– 58.02). Additionally, mortality was higher among PWH who were eligible but did not initiate TPT (aHR = 22.31; 95% CI: 4.47–111.35), as well as those who initiated but did not complete TPT (aHR = 13.75; 95% CI: 9.70–19.50), than those who completed TPT. Receiving care in referral hospitals showed a trend toward increased mortality compared to non-referral hospitals (aHR = 1.88; 95% CI: 0.96–3.64).

**Figure 2:**
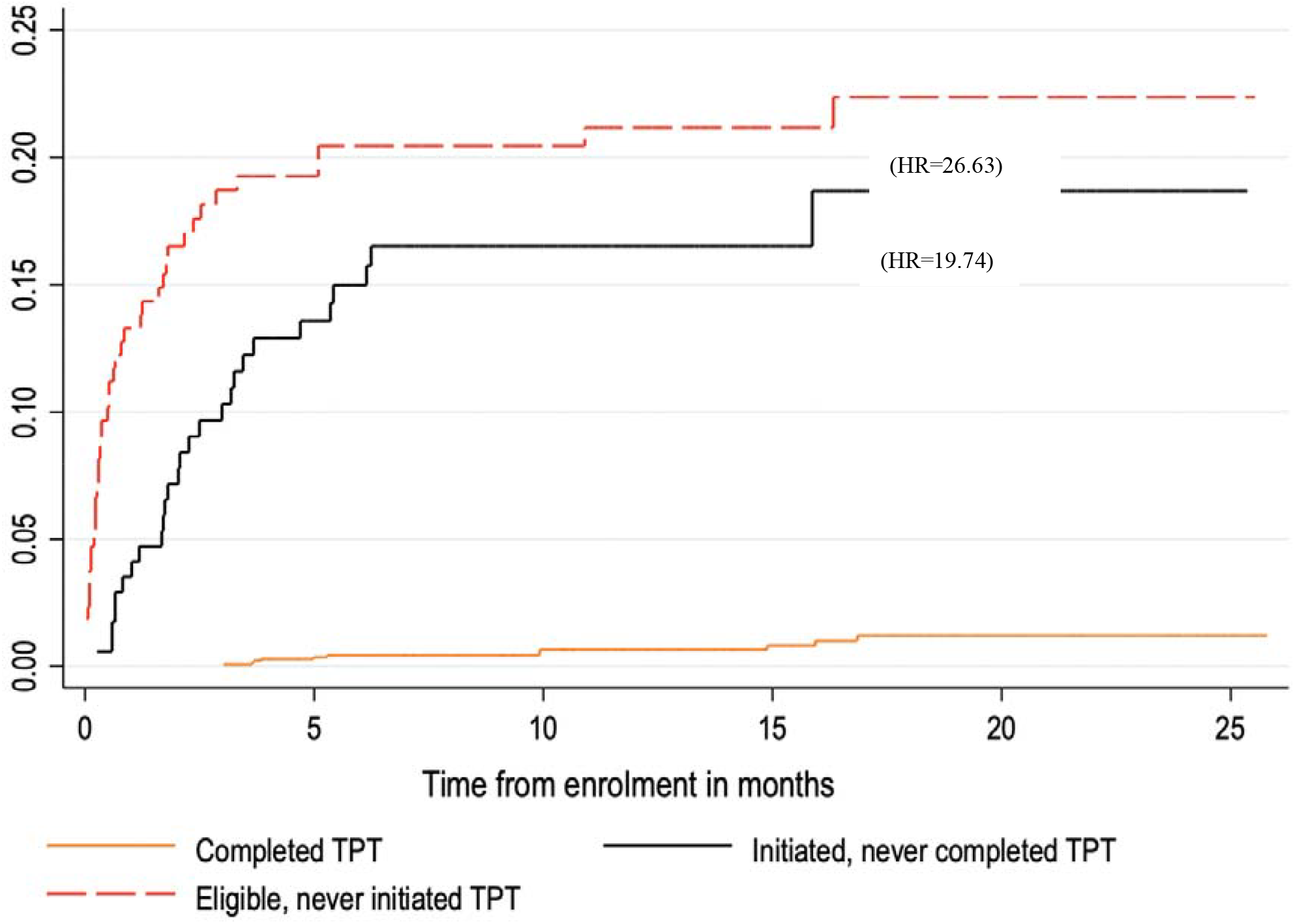
Nelson-Aalen curves depicting cumulative hazard of death within 24 months on enrolment in care among PWH who enrolled in care from January 2022 through September 2023, western Kenya (N=1, 930)

**Table 4:**
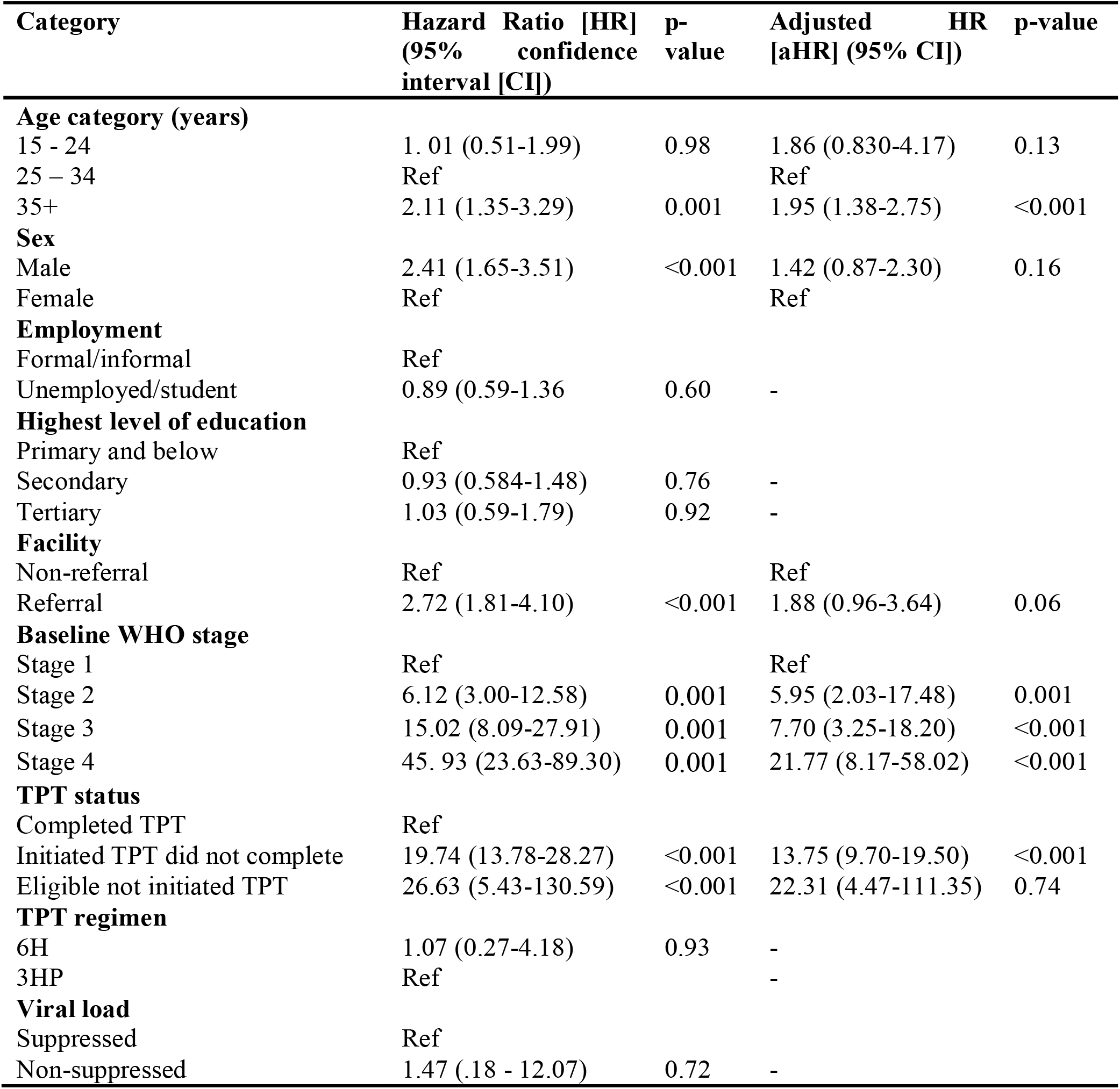
Risk factors for death among people with HIV who enrolled in HIV care from Jan 2022 through June 2023 in western Kenya (N=1, 930)

| Category | Hazard Ratio [HR]<br>(95% confidence interval [CI]) | p-value | Adjusted HR<br>[aHR] (95% CI) | p-value |
| --- | --- | --- | --- | --- |
| **Age category (years)** |  |  |  |  |
| 15 - 24 | 1.01 (0.51-1.99) | 0.98 | 1.86 (0.830-4.17) | 0.13 |
| 25 – 34 | Ref |  | Ref |  |
| 35+ | 2.11 (1.35-3.29) | 0.001 | 1.95 (1.38-2.75) | <0.001 |
| **Sex** |  |  |  |  |
| Male | 2.41 (1.65-3.51) | <0.001 | 1.42 (0.87-2.30) | 0.16 |
| Female | Ref |  | Ref |  |
| **Employment** |  |  |  |  |
| Formal/informal | Ref |  |  |  |
| Unemployed/student | 0.89 (0.59-1.36) | 0.60 | - |  |
| **Highest level of education** |  |  |  |  |
| Primary and below | Ref |  |  |  |
| Secondary | 0.93 (0.584-1.48) | 0.76 | - |  |
| Tertiary | 1.03 (0.59-1.79) | 0.92 | - |  |
| **Facility** |  |  |  |  |
| Non-referral | Ref |  | Ref |  |
| Referral | 2.72 (1.81-4.10) | <0.001 | 1.88 (0.96-3.64) | 0.06 |
| **Baseline WHO stage** |  |  |  |  |
| Stage 1 | Ref |  | Ref |  |
| Stage 2 | 6.12 (3.00-12.58) | 0.001 | 5.95 (2.03-17.48) | 0.001 |
| Stage 3 | 15.02 (8.09-27.91) | 0.001 | 7.70 (3.25-18.20) | <0.001 |
| Stage 4 | 45.93 (23.63-89.30) | 0.001 | 21.77 (8.17-58.02) | <0.001 |
| **TPT status** |  |  |  |  |
| Completed TPT | Ref |  |  |  |
| Initiated TPT did not complete | 19.74 (13.78-28.27) | <0.001 | 13.75 (9.70-19.50) | <0.001 |
| Eligible not initiated TPT | 26.63 (5.43-130.59) | <0.001 | 22.31 (4.47-111.35) | 0.74 |
| **TPT regimen** |  |  |  |  |
| 6H | 1.07 (0.27-4.18) | 0.93 | - |  |
| 3HP | Ref |  | - |  |
| **Viral load** |  |  |  |  |
| Suppressed | Ref |  |  |  |
| Non-suppressed | 1.47 (.18 - 12.07) | 0.72 | - |  |

## DISCUSSION

Our retrospective analysis of routine medical records of PWH newly enrolled in HIV care in western Kenya is among the first of its kind to demonstrate the impact of 3HP implementation on TPT initiation and completion compared to 6H in program settings. During the study period there was near-universal screening for TPT eligibility and almost 90% of TPT-eligible clients initiated 3HP, a higher rate of initiation than for 6H (84.2%). However, our analysis showed substantial non-completion of both 3HP and 6H regimens. Encouragingly, almost 90% of PWH in both study periods completed either 3HP or 6H, much higher completion rates than seen in prior programmatic data (20). 3HP completion was similar to 6H perhaps due to the generally high completion rates. This differs from clinical trials where completion of directly observed (DOT) 3HP exceeded 6H by up to 25% (5). Better adherence and completion of 3HP is largely attributed to its shorter duration (12 weekly doses) compared to 6H’s 6-month course (180 daily doses), substantially reducing the pill burden. While the completion of self-administered 3HP without reminders was non-inferior to DOT-administered 3HP in a randomized trial (21), our evaluation using real-world data suggest the need for additional adherence support measures to attain completion rates over 90%.

We found that low literacy, unemployment, receiving care in referral hospitals, and enrollment with AHD were associated with failure to initiate TPT. Lower literacy levels have been associated with poor TB-related health literacy and treatment uptake, including TPT (22). Educational disparities may impact health-seeking behavior and the ability to understand complex preventive treatment information. In addition, financial instability among the unemployed could limit access to healthcare and act as a barrier to TPT uptake. Previous studies have shown that larger hospitals had lower 6H initiation and completion rates than lower-level facilities (23, 24). Despite their advanced infrastructure, larger hospitals often struggle with complex service delivery models, leading to inefficiencies in TPT delivery (25). Decentralizing HIV care to lower-level facilities and providing additional support for referral hospitals may improve TPT uptake and completion as has been shown with ART (26, 27). Community-based models for TPT eligibility screening and delivery could further improve completion (28). Referral hospitals could also benefit from systems engineering tools, such as the systems analysis and improvement approach (SAIA), to optimize the TPT cascade (29). Nearly one in every five PWH were enrolled with AHD, reflecting delays in their HIV diagnosis and enrolment into care. Regrettably, PWH with AHD at enrolment had significantly reduced likelihood of initiating and completing both regimens. The lower TPT initiation among PWH with AHD highlights the challenges faced in delivering TPT to sicker PWH, probably due to a complex interplay of clinical, systemic, and patient-related factors. Higher rates of concurrent opportunistic infections among PWH with AHD may complicate TPT eligibility screening and lead to prioritization of other treatment needs over TPT (30).

In our study, 6H non-completion was higher in widowed or divorced PWH, those receiving HIV care in referral hospitals, and those with AHD at enrolment. Similarly, 3HP non-completion was also associated with being widowed or divorced and AHD at enrollment. The increased risk of TPT non-completion among widowed or divorced PWH could reflect reduced social support, which has been shown to have an impact on adherence to TB and HIV care (31). Strengthening social support mechanisms, such as peer-support programs, may help address adherence challenges among widowed or divorced individuals. Lower TPT completion rates in PWH with AHD could be due to worse adherence due to multiple medications, immune reconstitution inflammatory syndrome (IRIS), and deaths. Trials achieved over 90% 3HP completion rate in PWH under DOT, including those with AHD, suggesting that intensive support can mitigate barriers (5, 32).

Although retention in HIV care was high, all-cause mortality remained substantial. Mortality was strongly associated with AHD, probably due to susceptibility to opportunistic infections including tuberculosis, severe bacterial infections, and cryptococcal meningitis, which are leading causes of death in people with AHD (33). Prophylactic regimens, including TPT, are recognized to effectively reduce mortality in people with AHD (33), making it particularly concerning that TPT-eligible PWH with AHD were less likely to initiate and complete TPT. The similarly high mortality rates among those who were eligible but did not initiate TPT and those who did not complete TPT could indicate that incomplete therapy has limited protection. However, some of these deaths could have resulted from undiagnosed TB, which would not have been effectively treated with the preventive regimen. Improving TPT uptake and completion among PWH with AHD is critical, as the prevalence of AHD among PWH enrolling in HIV care has remained steady over time (34-36) due to delayed HIV diagnosis or default from ART, now recognized as a leading contributor to AHD at enrolment (35, 36). These findings underscore the importance of strengthening adherence support mechanisms to improve the completion of TPT regimens, including 3HP, among PWH with AHD.

Strengths of the study include data from high-volume HIV clinics in a region with a substantial HIV and TB burden providing valuable insights into the programmatic roll-out of shorter TPT regimens among PWH. However, routine program data is prone to incompleteness, which was minimized by abstracting data from multiple sources, including electronic medical records and hard copy registers. While we achieved completeness for most of the assessed variables, viral load and CD4 count information remained missing for over 40% of participants. This limited our ability to evaluate the role of viral load as a correlate of TPT non-initiation, TPT non-completion, and mortality. Because viral load testing is routinely performed three to six months after enrolment, only PWH who survived long enough after enrolment would have had their viral load assessed. This may have led to underestimation of the association of viral load non-suppression with mortality. Although the current Kenya ART guidelines recommend baseline CD4 testing to facilitate identification of PWH with AHD, a substantial proportion of participants in this study lacked documentation of CD4 counts. Consequently, we primarily relied on WHO clinical staging to identify PWH with AHD. This may have under-estimated the prevalence of AHD. This is supported by evidence from studies that have demonstrated that using CD4 count results in a higher prevalence of AHD (34-36).

In conclusion, this study demonstrated that 3HP implementation significantly increased TPT initiation while completion rates were high and similar for 3HP and 6H. TPT non-completion was substantially higher among PWH with AHD for both regimens. Mortality within 24 months of enrolment was substantial and higher among PWH with AHD and those who did not complete their TPT regimens. Targeted interventions are needed to support 3HP initiation and completion among PWH with AHD.

## Data Availability

Data are available upon request to the principal investigator.

## AUTHORSHIP

DOO, JK, GJ-S, and SML contributed to the conception, design, and execution of this study. DOO performed the analyses and drafted the initial manuscript. JM, LNN, SA, and TM critically reviewed the paper for analysis and interpretation of the data. All authors revised the manuscript critically and contributed to the final draft and read and approved the final manuscript.

## ACKNOWLEDGMENTS

We wish to acknowledge the staff who work in the HIV clinics who facilitated the implementation of this study. We also thank the research assistants who diligently abstracted data from the medical records of people with HIV whose information is presented in this paper. We further wish to extend our heartfelt gratitude to all the caregivers and children who participated in our study. Their contribution has been invaluable in advancing our understanding of adherence to isoniazid preventive therapy among children with HIV.

---

**Figure S1:**
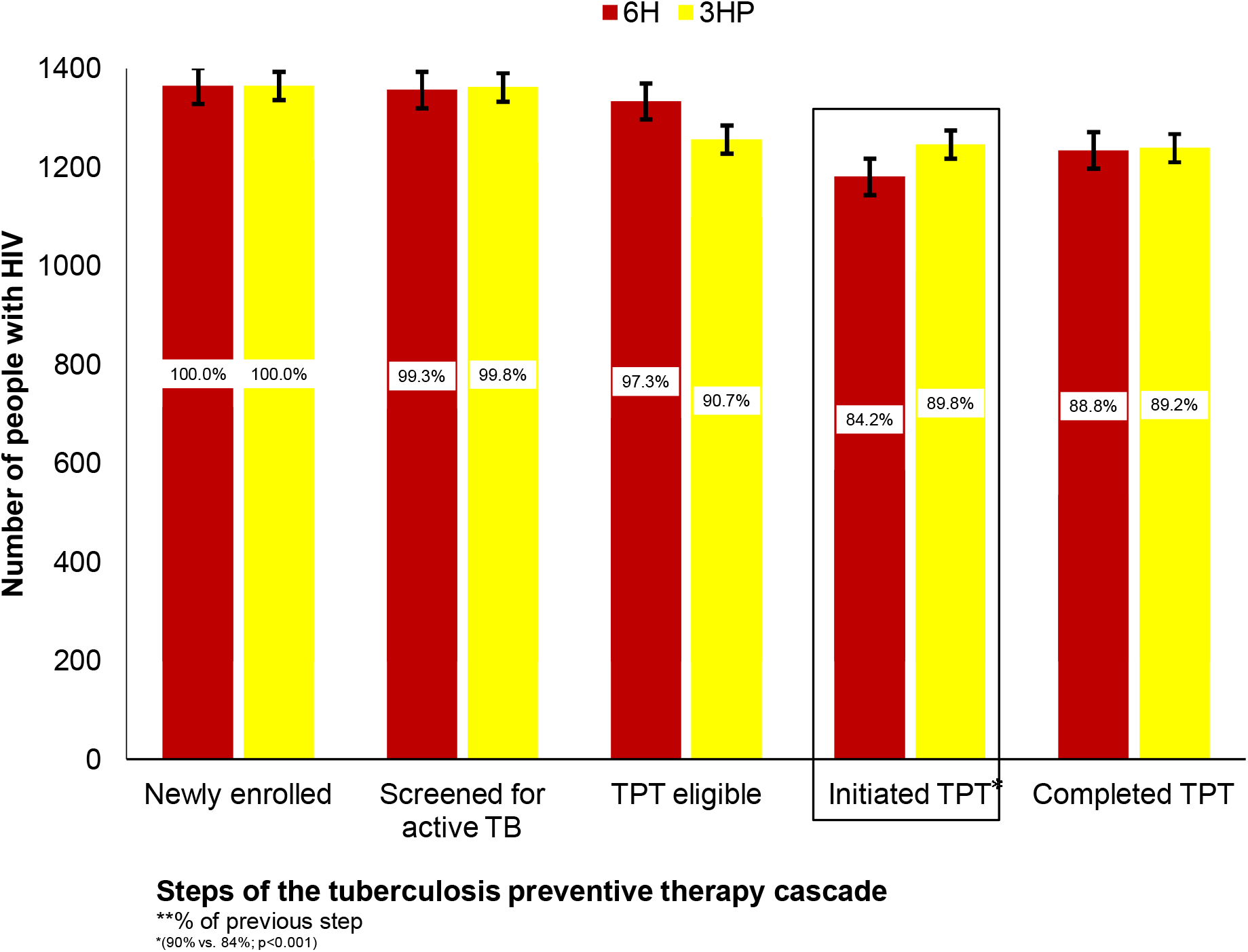
Comparison of six months of daily isoniazid (6H) vs. three months of weekly isoniazid with rifapentine (3HP) among people with HIV aged 15+, western Kenya, Jan 2022 to Sept 2023.

**Table S1:**
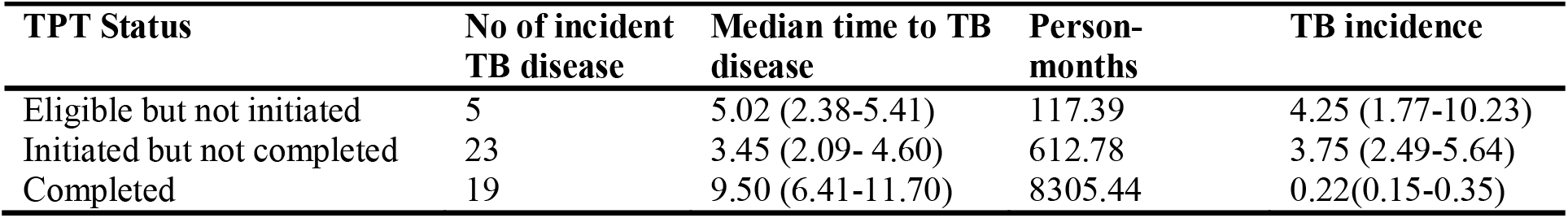
TB incidence rates among PWH who enrolled in care from January 2022 through September 2023, western Kenya (N=1, 930)

**Table S2:** Crude mortality rates among PWH who enrolled in care from January 2022 through September 2023, western Kenya (N=1, 930)

| **Category** | **Deaths** | **Person-years** | **Mortality rate/100 person years (95% CI)** |
| --- | --- | --- | --- |
| Overall | 108 | 21.7 | 5.0 (4.1-6.0) |
| **Age category (years)** |  |  |  |
| 15 - 24 | 12 | 3.6 | 3.3 (1.9-5.9) |
| 25 – 34 | 27 | 8.3 | 3.3 (2.2-4.8) |
| 35+ | 69 | 9.9 | 7.0 (5.5-8.9) |
| **Sex** |  |  |  |
| Male | 59 | 7.0 | 8.4 (6.5-10.9) |
| Female | 49 | 14.7 | 3.3 (2.5-4.4) |
| **Baseline WHO stage** |  |  |  |
| Stage 1 | 13 | 15.0 | 0.9 (0.50-1.48) |
| Stage 2 | 17 | 3.1 | 5.5 (3.4-8.8) |
| Stage 3 | 43 | 3.0 | 14.4 (10.7-19.4) |
| Stage 4 | 26 | 0.6 | 44.7 (30.4-65.6) |
| **TPT status** |  |  |  |
| TB disease at enrolment | 25 | 1.07 | 23.3 (15.8-34.5) |
| Eligible not initiated TPT | 44 | 2.3 | 18.8 (14.0-25.3) |
| Initiated TPT did not complete | 27 | 1.7 | 16.0 (10.9-23.3) |
| Completed TPT | 12 | 16.6 | 0.7 (0.4-1.2) |
| **TPT regimen** |  |  |  |
| 6H | 20 | 10.7 | 1.9 (1.2-2.9) |
| 3HP | 19 | 7.6 | 2.5 (1.6-2.9) |

